# Empirically calibrated allele frequency thresholds for ACMG BA1, BS1 and PM2 evidence criteria

**DOI:** 10.64898/2026.09.07.26362456

**Authors:** Viksar Dubey, Chris Eisenhart

## Abstract

Allele frequency (AF) is among the most frequently applied lines of evidence in variant classification, yet the ACMG/AMP criteria that use it (BA1, BS1, PM2) are still applied at fixed defaults while computational predictors have been systematically recalibrated. Population frequencies are shaped by selection, ascertainment, and gene-level demography at once, and few genes carry enough classified variants to set a threshold directly. Extending the calibration approach applied to computational predictors, we used inheritance mode and gene-level missense constraint as stratification axes and pooled variants within each stratum. ClinVar missense variants annotated against gnomAD v4.1.1 were stratified along both, and gene-normalized kernel density estimates were fit to pathogenic and benign variants within a sliding window along the constraint axis. Thresholds were placed where the likelihood ratio crossed ACMG/AMP evidence strengths at a prior of 0.0441. Derived thresholds varied systematically with constraint and differed between inheritance modes, departing from the fixed defaults in both directions. On held-out genes, stratified cutoffs reached 96.7% accuracy against 90.1% unstratified. Restricted to the 73 ClinGen expert panel genes with autosomal dominant or recessive inheritance, the derived cutoffs reached 91.0% accuracy at 69.5% variant coverage, against 88.8% accuracy at 86.2% coverage for the panel-specified cutoffs. AF thresholds for these criteria are not constant across genes, and inheritance mode and missense constraint capture much of that variation. The resulting cutoffs are empirically derived, carry explicit uncertainty, and deploy as a lookup table across thousands of genes no expert panel currently covers.

## 1. Introduction

The ACMG/AMP sequence variant interpretation guidelines assign population allele frequency (AF) a central role in classification^1^. Three criteria rest on it directly. BA1 marks a variant as benign on frequency alone, without recourse to any other line of evidence; BS1 provides strong benign evidence when a variant is more common than the disorder could plausibly support; PM2 credits absence or extreme rarity as evidence toward pathogenicity^1^.

The thresholds attached to these criteria have not kept pace with the framework around them. The Bayesian reformulation of the guidelines placed every criterion on a common logarithmic scale, expressing evidence strength as an odds ratio and making explicit what each criterion is worth^2^. Computational predictors were subsequently recalibrated against that scale, first for thirteen tools and later for a further three, with thresholds derived from local posterior probabilities rather than developer recommendations^3,4^. Frequency evidence received no comparable treatment. In most laboratories BA1 and BS1 are still applied at 5% and 1%, values that entered practice as expert consensus rather than derived from data^1^. The ClinGen Sequence Variant Interpretation Working Group excluded predictors that incorporate AF from its calibration efforts on two grounds: a tool with frequency built into it cannot be combined with BA1 or BS1 without counting the same evidence twice, and calibrating such tools across frequency strata would require more classified variants than are currently available^4^. Frequency evidence, in other words, has been deliberately quarantined so that it can be applied on its own. It has not yet been calibrated for that purpose.

There is a further reason frequency resists the treatment given to predictors. A prediction score is a single number per variant, comparable across the genome by construction, while an AF is not. The same 0.5% AF carries different weight in a constrained dominant gene than in a tolerant recessive one, because the distribution of frequencies a gene can sustain depends on selection acting through its mode of inheritance, on the demographic history of the reference population, and on which variants happened to be submitted for classification^5^. Per-gene counts compound the problem. Most genes carry too few classified variants for a threshold to be estimated from that gene alone. The established response to this has been to model the disease rather than the data. Whiffin and colleagues derive a maximum credible population allele frequency from disease prevalence, allelic and genetic heterogeneity, inheritance mode, and penetrance, together with the sampling variance of the reference dataset^5^. The approach is principled and is now embedded in practice guidelines, which recommend it for setting BA1 and BS1 on a per-gene basis^6^. Its constraint is the input it requires. Reliable prevalence and penetrance estimates are unavailable for most genes, and groups applying the method have reported exactly this difficulty, resorting to conservative bounds where the parameters could not be pinned down^6^. Expert panels address the same problem gene by gene, curating thresholds with per-gene evidence and sometimes departing from the defaults by more than an order of magnitude^7^, though this per-gene curation has been completed for only a small fraction of clinically interpreted genes.

We extend the calibration approach of Pejaver and Bergquist^3,4^, who set predictor thresholds at the Bayesian evidence strengths, to the AF-based criteria the same working group had to exclude from their scope. The unit being calibrated is an allele frequency rather than a predictor score, so a single threshold across the genome will not do. Instead we stratify along the two axes most likely to shift the pathogenic/benign frequency distribution: mode of inheritance, which separates the two selective regimes most likely to differ, and gene-level missense constraint^8^, which orders genes within each regime by how much variation they tolerate. Pooling variants within strata gives each cell enough classified data for direct estimation, extending coverage to the genes for which the per-gene prevalence and penetrance inputs Whiffin–Ware requires are not currently available. This yields empirically derived AF thresholds for BA1, BS1, and PM2 across 4,527 genes, each accompanied by a bootstrap confidence band, discretized into a lookup table. Where expert panel specifications exist we compare against them directly. Where they do not, which is most genes, these cutoffs provide an alternative to a fixed default.

## 2. Material and Methods

### 2.1. Variant Dataset

The full ClinVar VCF^9^ was downloaded from the NCBI FTP site in August 2026. The release contained 4,461,717 variants. The full filtering flow from this release to the final calibration set is shown in Figure 1. We opted to retain only missense variants, whose AF distributions reflect selective pressures that differ systematically from loss-of-function (LoF) variants^10^ and would not be expected to share similar frequency thresholds. Among all missense variants, we retained those with an assertion of pathogenic, likely pathogenic, benign, or likely benign^1^ and at least one-star review status. Variants of uncertain significance and those with conflicting interpretations fell out under the former criterion; zero-star variants, under the latter. These filters left 207,907 variants across 15,022 genes. These variants were then annotated with Nirvana^11^, yielding population allele frequencies (AFs) from gnomAD v4.1.1^8^. AF was taken as the maximum frequency observed across gnomAD’s continental subpopulations (popmax) rather than the pooled global AF, guarding against a variant that looks rare overall but is in fact common within a single population^5^. The remaining variants were assigned a mode of inheritance (MOI) based on their respective gene, drawn from HPO^12^, with GenCC^13^ and ClinGen^14^ as fallbacks. Gene-level missense constraint was quantified using the missense observed/expected ratio (Missense O/E) and its upper confidence bound metric^10^, obtained from gnomAD v4.1.1 constraint metrics^8^, with MANE Select transcripts^15^ preferred where more than one was available. We used the upper bound rather than the point estimate because in shorter genes, where the expected variant count is small and sampling noise is large, a point estimate can understate constraint. The upper bound is the more conservative choice, and it follows the same convention gnomAD uses for LOEUF, its widely adopted loss-of-function constraint metric^10^. A further 1,490 variants across 209 genes had no constraint data and were dropped here. Calibration was restricted to genes with purely autosomal dominant (AD) or autosomal recessive (AR) inheritance patterns. Genes with an unknown MOI, both AD and AR inheritance patterns, or X-linked inheritance patterns were excluded from this analysis, removing 93,827 variants and leaving 112,590 across 4,609 genes. Additionally, at this point in the filtration process, 31,663 of the 112,590 ClinVar variants were absent from gnomAD and therefore lacked population-level AF. These were excluded rather than assigned a pseudocount frequency, leaving 36,980 AD and 43,947 AR variants across 4,527 genes. The exclusion fell disproportionately on pathogenic variants (64.5%, versus 10.0% of benign), which overall aligns with the fact that absence from gnomAD is a result of rarity.

**Figure 1:**
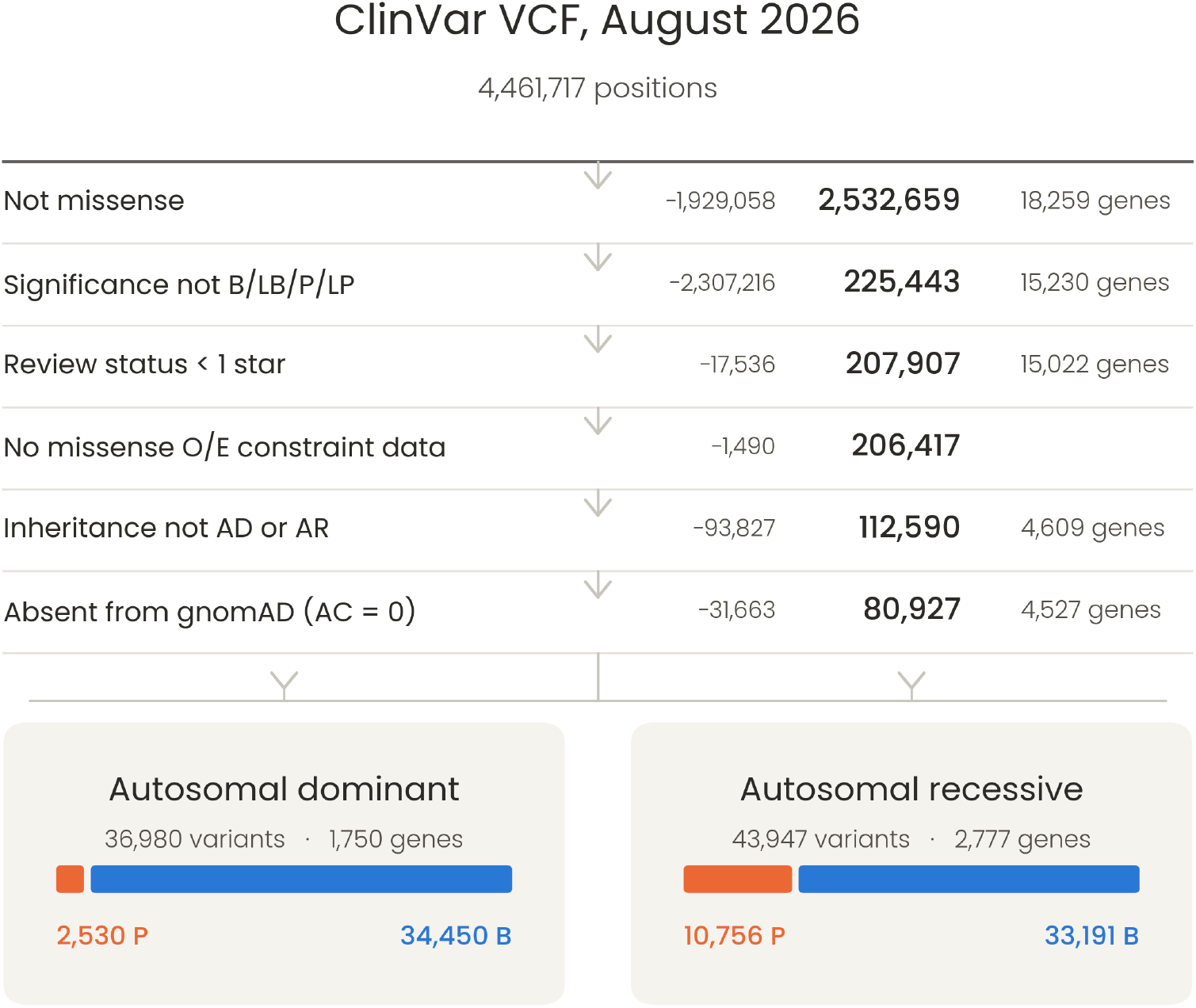
Filtering flow from the ClinVar release to the final calibration set. Gene counts are unavailable before the missense filter, since gene assignment follows annotation.

### 2.2. Cutoff Derivation

Cutoffs were derived independently for each MOI. Rather than binning genes into fixed constraint intervals, we slid a window along the missense O/E upper confidence bound axis in steps of 0.005, spanning 0.15 to 1.80. At each center point, the window was required to contain at least 300 pathogenic and 300 benign variants drawn from no fewer than 15 distinct genes. Near the ends of the axis, where less data is available on one side, these minimums were scaled in proportion to the overlap^3^. This keeps the estimate stable in sparse regions without over-smoothing the dense ones. Within each window, we fit a Gaussian kernel density estimate on log^10^(AF), separately for benign and pathogenic variants, using a fixed bandwidth of 0.03. Each variant was weighted by the inverse of its gene’s variant count, so that a small number of heavily sequenced genes could not dominate the local density. For a class *c ∈ {P, B}* and a window containing variants *i* in genes *g*(*i*),

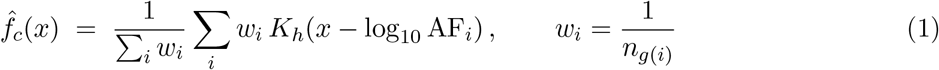

where *K*_*h*_ is a Gaussian kernel with *h* = 0.03 and *n*_*g*_ is the number of variants in gene *g*. The likelihood ratio at a given AF is then

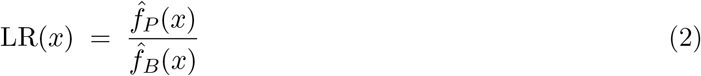

Thresholds were placed where this likelihood ratio crosses the targets specified by the Bayesian adaptation of the ACMG/AMP framework^2^. Under that framework, an evidence constant *C* is defined so that combinations of criteria reproduce the ACMG/AMP classification rules at a given prior^2^. We adopted a prior probability of pathogenicity of 0.0441^3^, which yields *C* = 1124 and evidence strengths

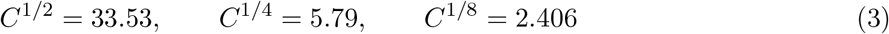

for strong, moderate, and supporting respectively. BA1 is stand-alone under ACMG/AMP rather than a strength tier^1^. We mapped it to the very strong value of *C*, which is both the conventional choice and the conservative one. Uncertainty was quantified by cluster bootstrap. Because variants within a gene are not independent, we resampled genes rather than variants; resampling variants individually would understate the variance. Each of 400 iterations drew a full set of genes with replacement, refit the densities, and re-derived every threshold. The reported 95% confidence bands are the 2.5th and 97.5th percentiles across those iterations. The resulting cutoff-versus-constraint curves were smoothed with a Gaussian kernel of bandwidth 0.10. For deployment, we discretized the smooth curves into bins 0.1 units wide and took the median cutoff within each bin, trimming edge bins that contained fewer than 15 genes. This yields a step-function lookup table for each MOI.

### 2.3. Evaluation Metrics

Accuracy is computed only among variants that received a call. A variant is judged if its AF crosses either the PM2_Supporting threshold on the low side or the BS1_Supporting threshold on the high side. Benign variants count as correct under any benign tier, pathogenic variants under PM2_Supporting. Anything falling between the two thresholds is left indeterminate and does not enter the calculation. Errors were also scored by the strength with which they were made. Each tier carries a signed evidence weight on the log^2^ LR scale^2^, running from *−*10.13 bits at BA1 through *−*5.07, *−*2.53, and *−*1.27 for the weaker benign tiers, 0 for an indeterminate call, and +1.27 for PM2_Supporting. For a variant with true label *y ∈ {*+1, *−*1*}* and assigned weight *s*,

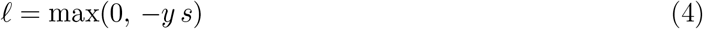

Correct calls contribute nothing, since the signs agree. Wrong calls cost the full weight of the tier assigned, so calling a pathogenic variant BA1 costs 10.13 bits while calling it BS1_Supporting costs 1.27. The benign side spans four tiers reaching *−* 10.13 bits while the pathogenic side has only PM2_Supporting at +1.27, which reflects the clinical reality that the two errors are not equally bad. Loss is averaged over all variants and reported in bits per variant. Variants reaching no threshold contribute zero, since under ACMG/AMP the correct output for insufficient evidence is no claim rather than a forced one^1^.

### 2.4. Held-out Gene Validation

Cutoffs were evaluated on a held-out set of genes. We split genes 90/10 rather than splitting variants, since variants within a gene are not independent and a variant-level split would leak information across the boundary. The test set was class-balanced by downsampling the majority class within each MOI, giving 3,074 variants (456 AD, 2,618 AR) across 372 genes (105 AD, 267 AR; no gene-level overlap between the two MOI subsets).

Four approaches were compared, each adding one layer of stratification. The first serves as a control: a single set of AF cutoffs applied to every variant regardless of inheritance mode or constraint, derived from one KDE fit on all AD and AR variants pooled. The second isolates the contribution of MOI alone. It fits one KDE per inheritance mode, giving separate cutoffs for AD and AR, but leaves the constraint axis untouched. The third is the deployment-ready version and the one we intend for use. The constraint axis is divided into bins 0.1 units wide, each bin carrying its own cutoffs read off the smooth curves, so that classifying a variant amounts to finding the bin matching its gene’s missense O/E upper confidence bound and reading the values. The fourth is the smooth curve itself, interpolated at the exact constraint value rather than binned. It is what the sliding window produces before discretization, and comparing it against the binned version shows whether discretization costs any meaningful resolution. Comparing the first two isolates the effect of MOI stratification; comparing the second against the third isolates the effect of constraint stratification; comparing the third against the fourth measures the cost of discretization.

### 2.5. Comparison against VCEP Cutoffs

We compared our derived cutoffs against those published by ClinGen Variant Curation Expert Panels^14^. Specification documents GN002 through GN243 were scraped from the CSpec Registry, and we collected gene-level BA1, BS1, BS1_Moderate, BS1_Supporting, and PM2_Supporting thresholds from the 114 released specifications that stated them explicitly, covering 132 genes across 42 expert panels. Values expressed as percentages were converted to fractions, and thresholds embedded in methodological preamble (e.g. case frequencies, penetrance estimates) were excluded using context-aware filtering. Where panels publish gene-specific thresholds, these can depart from the ClinGen defaults by more than an order of magnitude^7^. Genes were matched to our calibration dataset by exact HGNC symbol. Of the 132 matched genes, 79 could be assigned an unambiguous autosomal dominant or autosomal recessive mode of inheritance; the remaining 53 (mixed, X-linked, or unresolved) were excluded for the same reason such genes were excluded from calibration. Nine of the 79 genes carry both AD and AR VCEP entries and so contribute to both inheritance strata in the comparison that follows (54 AD, 34 AR). Of the 79, 73 (46 AD, 27 AR) have benign or pathogenic ClinVar missense variants in the calibration dataset and enter the downstream classification comparison.

## 3. Results

### 3.1. Derived Cutoffs

The derived cutoffs for each MOI are given in Tables 1 and 2. Both climb with missense O/E, though not at the same pace or in the same shape. AD moves in a near-monotonic staircase, while AR climbs more unevenly and sits several-fold above AD at comparable constraint. PM2_Supporting barely moves across the range in either mode. Both tables start above the sliding window’s nominal 0.15 lower bound (Methods 2.2), since bins with fewer than 15 genes were trimmed for insufficient data. AD’s deployed range begins at missense O/E 0.4 and AR’s range begins at 0.7.

**Table 1:** Derived AD cutoffs by missense O/E upper confidence bound bin.

| Bin | BA1 | BS1 | BS1_Mod | BS1_Sup | PM2_Sup |
| --- | --- | --- | --- | --- | --- |
| [0.4, 0.5) | 1.906e-03 | 1.605e-03 | 4.356e-04 | 4.068e-04 | 3.843e-06 |
| [0.5, 0.6) | 2.490e-03 | 2.106e-03 | 8.695e-04 | 7.658e-04 | 4.137e-06 |
| [0.6, 0.7) | 3.545e-03 | 3.048e-03 | 1.757e-03 | 1.268e-03 | 4.328e-06 |
| [0.7, 0.8) | 5.255e-03 | 4.337e-03 | 2.468e-03 | 1.222e-03 | 4.616e-06 |
| [0.8, 0.9) | 7.961e-03 | 5.979e-03 | 3.399e-03 | 1.186e-03 | 5.052e-06 |
| [0.9, 1.0) | 9.619e-03 | 7.401e-03 | 5.021e-03 | 1.531e-03 | 5.426e-06 |
| [1.0, 1.1) | 9.694e-03 | 8.354e-03 | 6.694e-03 | 2.678e-03 | 6.059e-06 |
| [1.1, 1.2) | 9.871e-03 | 9.004e-03 | 8.014e-03 | 5.280e-03 | 6.550e-06 |
| [1.2, 1.3) | 1.004e-02 | 9.249e-03 | 8.638e-03 | 7.559e-03 | 6.486e-06 |
| [1.3, 1.4) | 1.007e-02 | 9.287e-03 | 8.797e-03 | 8.193e-03 | 6.108e-06 |
| [1.4, 1.5) | 1.007e-02 | 9.289e-03 | 8.838e-03 | 8.282e-03 | 5.829e-06 |

**Table 2:**
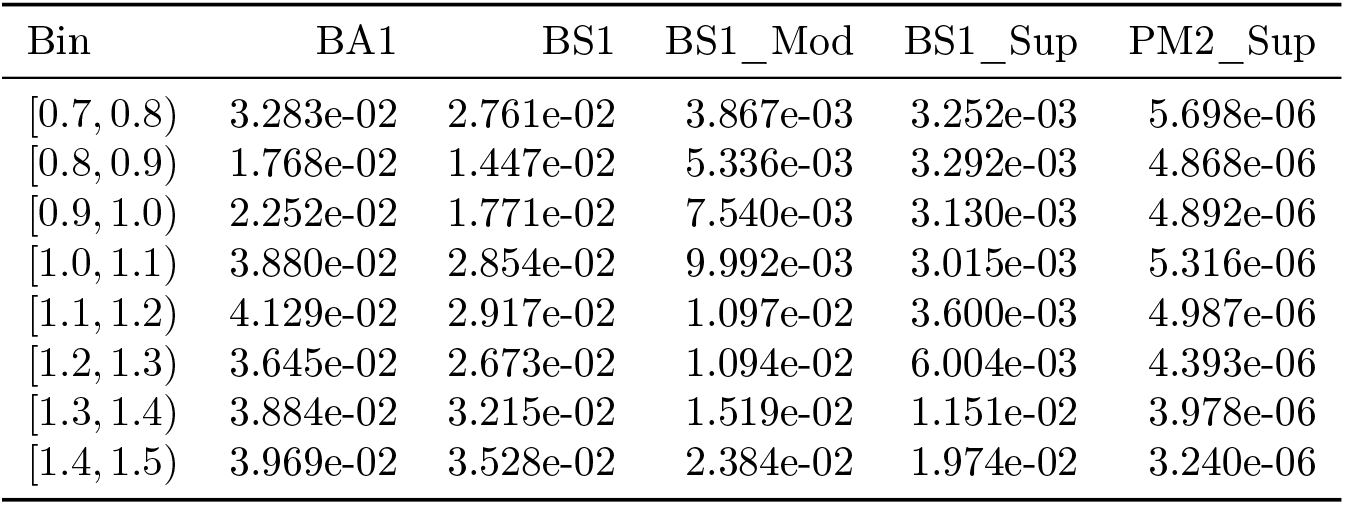
Derived AR cutoffs by missense O/E upper confidence bound bin.

| Bin | BA1 | BS1 | BS1_Mod | BS1_Sup | PM2_Sup |
| --- | --- | --- | --- | --- | --- |
| [0.7, 0.8) | 3.283e-02 | 2.761e-02 | 3.867e-03 | 3.252e-03 | 5.698e-06 |
| [0.8, 0.9) | 1.768e-02 | 1.447e-02 | 5.336e-03 | 3.292e-03 | 4.868e-06 |
| [0.9, 1.0) | 2.252e-02 | 1.771e-02 | 7.540e-03 | 3.130e-03 | 4.892e-06 |
| [1.0, 1.1) | 3.880e-02 | 2.854e-02 | 9.992e-03 | 3.015e-03 | 5.316e-06 |
| [1.1, 1.2) | 4.129e-02 | 2.917e-02 | 1.097e-02 | 3.600e-03 | 4.987e-06 |
| [1.2, 1.3) | 3.645e-02 | 2.673e-02 | 1.094e-02 | 6.004e-03 | 4.393e-06 |
| [1.3, 1.4) | 3.884e-02 | 3.215e-02 | 1.519e-02 | 1.151e-02 | 3.978e-06 |
| [1.4, 1.5) | 3.969e-02 | 3.528e-02 | 2.384e-02 | 1.974e-02 | 3.240e-06 |

The reliability of these estimates varies across the constraint axis, and the bootstrap in Methods 2.2 was designed to quantify exactly that. Figure 2 plots the smooth cutoff-versus-constraint curve behind Tables 1 and 2, point estimate in black, together with its 95% confidence band from 400 gene-resampling bootstrap iterations. Grey shading marks the stretch of the constraint axis outside the deployed bin range; the tails where fewer than 15 genes fell inside the sliding window and the corresponding bins were trimmed from the lookup table, so the curve is shown there for completeness rather than for use. Within the deployed range, the bands are narrowest through the well-populated middle of the axis and widen toward its edges, most visibly for PM2_Supporting in both modes and for BS1_Moderate in AR at low constraint, exactly where gene counts are thinnest. Panel B shows the point estimate for BA1 in AR dipping and then recovering between constraint 0.7 and 1.0, the unevenness noted earlier, now bracketed by an interval wide enough to indicate that the dip reflects signal rather than noise.

**Figure 2:**
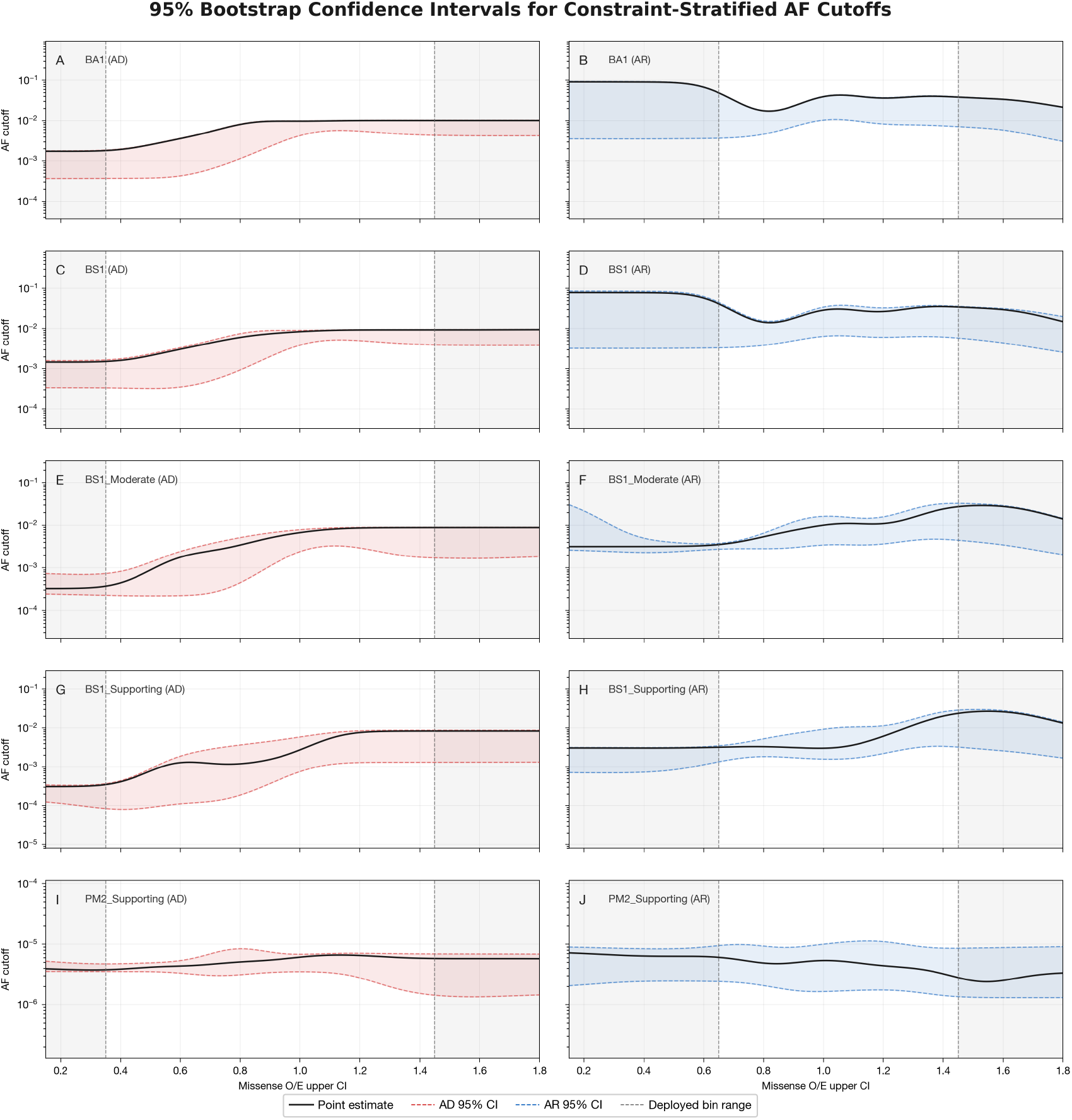
Point estimates (black) and 95% bootstrap confidence bands (400 gene-resampling iterations, 2.5th–97.5th percentiles; Methods 2.2) for the smooth cutoff-versus-constraint curve behind each criterion, shown separately for AD (red) and AR (blue). Deployed cutoffs (Tables 1, 2) are the median of this curve within each 0.1-unit bin; grey shading marks the constraint range outside those deployed bins, trimmed from the lookup table for containing fewer than 15 genes. Bands narrow through the well-populated middle of the constraint axis and widen toward its sparser edges.

### 3.2. Held-out Gene Validation

Model performance is summarized in Figure 3. The fully stratified model (per-MOI, constraint-stratified) reached 96.7% accuracy at 36.9% coverage, with a loss of 0.0173 bits per variant. The binned version used for deployment performed nearly identically: 96.6% accuracy, 36.5% coverage, 0.0169 bits. The per-MOI flat model reached 90.4% accuracy at 63.8% coverage (0.0804 bits), and the fully pooled model reached 90.1% accuracy at 64.2% coverage (0.0845 bits).

**Figure 3:**
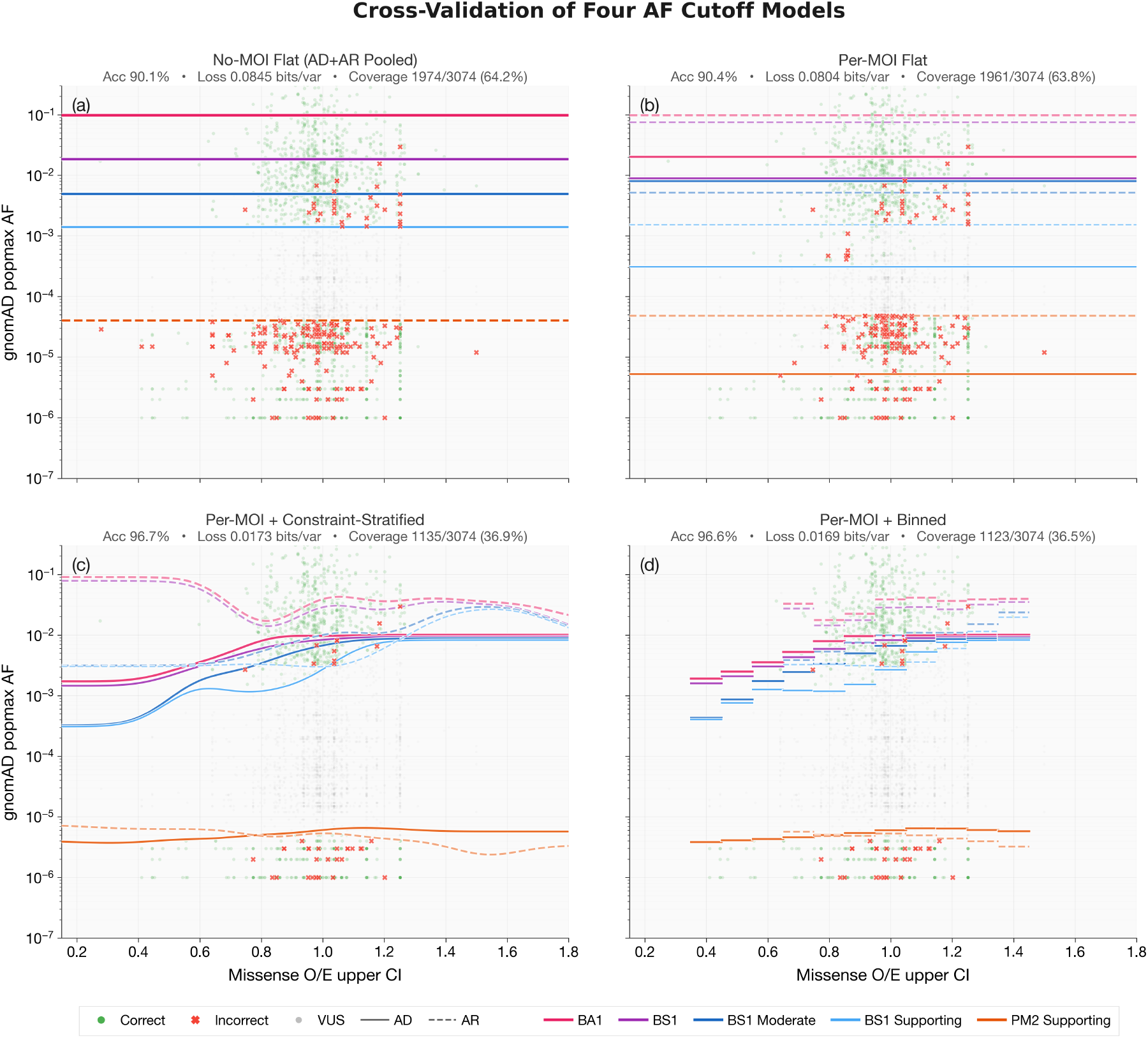
Four-model ablation on the held-out gene set. Panels progress from fully stratified (per-MOI + constraint-stratified, left) to fully pooled (no MOI, no constraint, right). Green points are correctly classified variants; red points are misclassified. Metrics shown per panel: accuracy among called variants, loss in bits per variant on the log_2_ likelihood ratio scale, and coverage as the fraction of variants receiving a firm call. Lower coverage in the stratified panels reflects a wider VUS zone by design; the cutoffs commit only where the empirical variant distribution supports a call.

Separating AD from AR without constraint stratification produced only a marginal gain over the pooled model. Constraint stratification within each mode improved accuracy from 92.0% to 95.3% in AD and from 90.2% to 96.8% in AR. Relative to the pooled model, the stratified cutoffs improved 158 held-out variants and worsened 693; most of the worsened calls were previously correct calls that became indeterminate, rather than calls that flipped to the wrong class.

### 3.3. Comparison against VCEP Cutoffs

VCEP-published cutoffs are the applied product of the Whiffin–Ware framework combined with expert-curated epidemiological inputs, so comparing against them shows where the stratified estimates agree with the per-gene principled approach and where they diverge, without requiring us to re-estimate the prevalence and penetrance parameters the panels have already curated. The two sets diverge in a systematic pattern (Figure 4). The derived step functions place BA1, BS1, BS1_Moderate, and BS1_Supporting at consistently higher allele frequencies than the VCEP-published gene-specific values, while placing PM2_Supporting at consistently lower frequencies. The consequence is a wider VUS zone under the derived cutoffs: variants at intermediate frequencies receive no AF-based call under our thresholds but are assigned a benign or pathogenic supporting code under the panel-specified ones.

**Figure 4:**
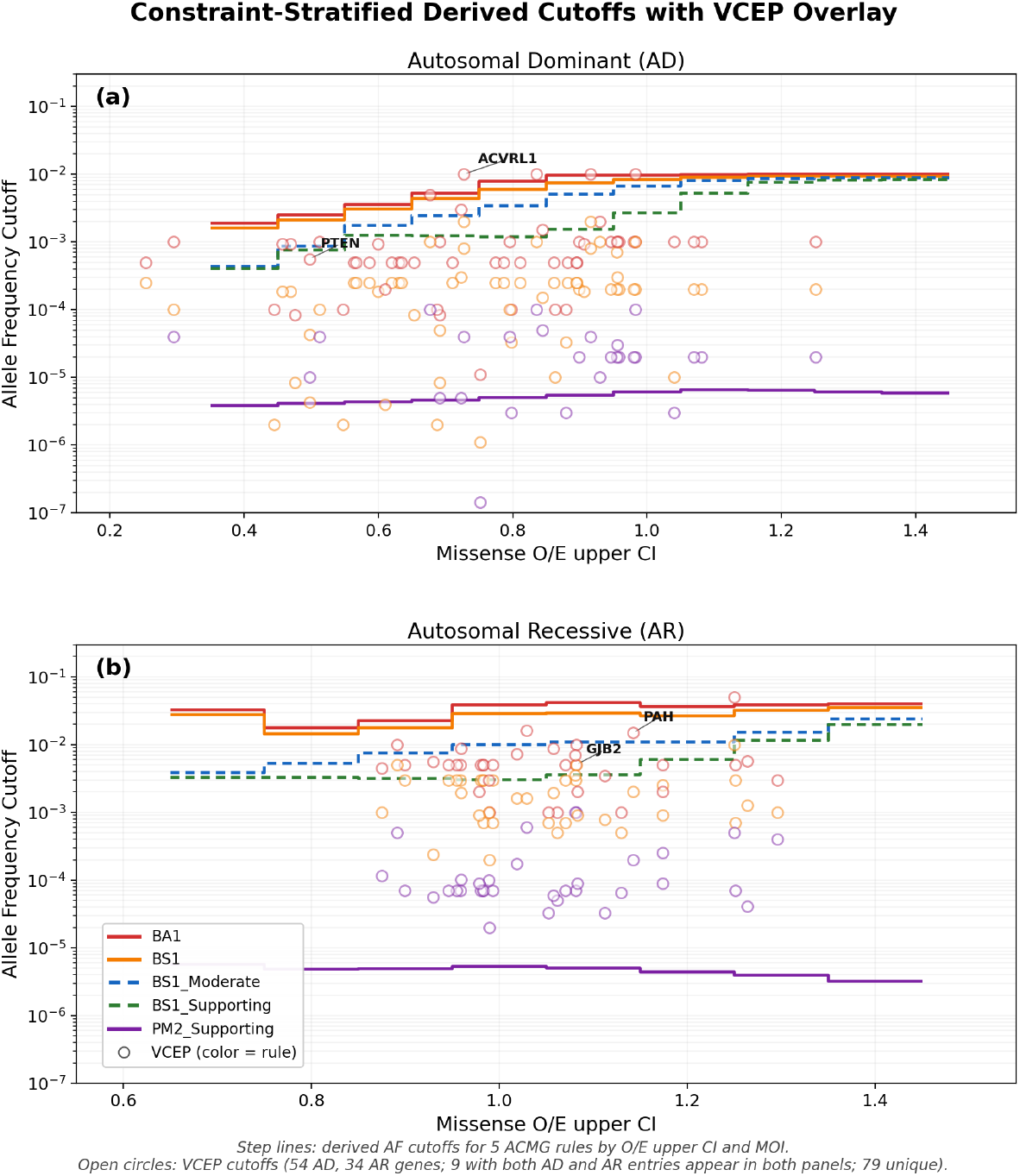
Derived AF cutoffs (step lines) with VCEP-published cutoffs (open circles) overlaid for the 88 matched genes with autosomal dominant or recessive inheritance (54 AD, 34 AR). Left: autosomal dominant. Right: autosomal recessive. The derived step functions sit above the VCEP dots for the benign tiers and below them for PM2_Supporting, producing a wider VUS zone. Of these 88 genes, 73 (46 AD, 27 AR) also have held-out variants and form the accuracy comparison reported in the remainder of this section.

Restricted to those 73 genes, the derived and panel-specified cutoffs again diverged in overall accuracy and in how many variants each was willing to call (Figure 5). Across all 11,260 variants, the derived cutoffs reached 91.0% accuracy at 69.5% coverage, against 88.8% accuracy at 86.2% coverage for the panel-specified cutoffs. Splitting by inheritance mode, the 46 AD genes (7,527 variants) showed close overall accuracy either way (90.6% derived versus 89.7% panel-specified) with derived coverage at 80.9% against the panels’ 88.2%; by class, derived pathogenic accuracy reached 100.0% against 98.0% for the panel-specified cutoffs, while derived benign accuracy was 34.8% against 57.1%. In the 27 AR genes (3,733 variants), derived accuracy was higher (92.4% against 86.9%) and derived coverage was lower still, at 46.6% against the panels’ 82.0%; derived benign accuracy was 75.1% against 58.0% and derived pathogenic accuracy was 98.7% against 98.3%.

**Figure 5:**
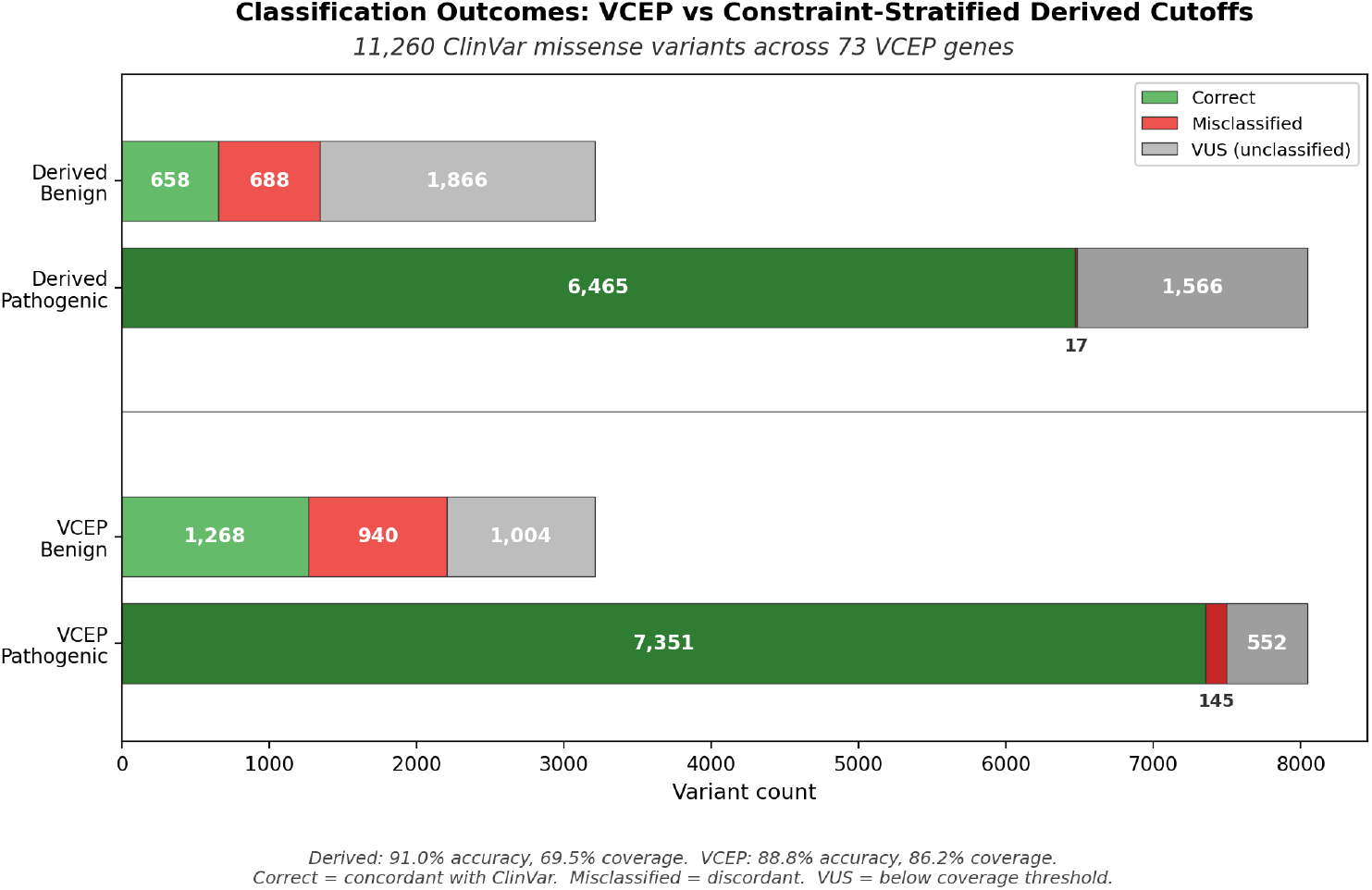
Classification outcomes across the 11,260 ClinVar variants in the 73 VCEP comparison genes restricted to autosomal dominant or recessive inheritance (46 AD, 27 AR). The derived cutoffs reach higher overall accuracy than the panel-specified cutoffs (91.0% versus 88.8%) at lower coverage (69.5% versus 86.2%), with the balance of pathogenic- and benign-side accuracy differing by inheritance mode (Results 3.3).

Two patterns emerge when AD and AR are compared directly. First, the size of the accuracy gap tracks the size of the coverage gap: in AD both are modest, with derived accuracy close to the panel-specified value and derived coverage only slightly lower; in AR both are larger, with derived accuracy exceeding the panels by a wider margin while derived coverage falls further below theirs. Second, the benign-side comparison reverses direction between modes: derived cutoffs underperform the panels on benign accuracy in AD but outperform them in AR, even as pathogenic-side accuracy remains comparable between the two cutoff sets in both modes.

## 4. Discussion

Several practical points bear on applying these cutoffs. A gene whose constraint falls outside the deployed bin range should be assigned the nearest deployed bin, or the ACMG/AMP default where the nearest bin is far enough from the observed constraint that borrowing from it strains the interpolation. Loss-of-function variants, indels, and genes with mixed, X-linked, or mitochondrial inheritance sit outside the calibrated scope. These should be classified against the ACMG/AMP defaults or an expert panel specification where one exists. Where a VCEP specification exists for a covered gene, the panel specification is the appropriate primary reference; the panels integrate gene-specific evidence far beyond what any general calibration can access. The derived cutoffs are intended as a complement, not a substitute, and function as an independent cross-check whose disagreements flag epidemiological inputs that may warrant re-examination. Consistent with the SVI 2020 downgrade of PM2 from Moderate to Supporting^16^, a conclusion many VCEPs had already reached independently, the pathogenic-side likelihood ratio does not reach the moderate evidence target in either inheritance mode; PM2 appears in the derived tables at Supporting only. The lookup tables are tied to the specific ClinVar and gnomAD releases used here and are expected to be re-fit as those datasets accumulate.

### 4.1. VUS zone

Lower coverage is a direct consequence of applying the Bayesian evidence framework to the empirical variant distribution. Thresholds fire only where the distribution supports a call, so variants at intermediate frequencies are handed to the remaining ACMG/AMP evidence lines rather than assigned an AF-based code on their own.

Our derived cutoffs place a distinct AF band between the BS1_Supporting and PM2_Supporting thresholds in which no AF-based criterion fires. A variant falling within this range receives no contribution from BA1, BS1, or PM2, and its classification rests on the remaining ACMG/AMP evidence lines. We refer to this band as the VUS zone with the caveat that a specific variant’s final classification depends on those other criteria, not on AF alone. The zone is present across every MOI and missense O/E stratum. It is wider than the band produced by the panel-specified cutoffs, because most expert panels apply BA1 and BS1 at their original 2015 ACMG/AMP strengths and have not adopted the SVI 2020 tier-downgrade framework^16^, so BS1_Supporting is not part of the panel-specified cutoff set. Adopting the tier-downgrade framework at the AF-based criteria is the same recalibration effort that was applied to computational predictors, and a wider VUS zone is its expected consequence: the AF criteria commit to a call only where the empirical variant distribution supports one, leaving borderline cases to be resolved by segregation, functional, or computational evidence.

### 4.2. Derived AF vs VCEP

The 42 ClinGen expert panels covered here represent a sustained curation effort over many years, integrating gene-specific evidence beyond what any general calibration can access. Our comparison is consistent with that expertise: pathogenic-side accuracy under the panel-specified cutoffs sits at 98% or above in both inheritance modes, matching or nearly matching the derived cutoffs, and the two approaches diverge principally at the benign-side boundaries. On the 73 ClinGen expert panel genes restricted to autosomal dominant or recessive inheritance, the derived cutoffs again committed to fewer calls than the panel-specified cutoffs (69.5% coverage against 86.2%), but the accuracy comparison no longer moves in one direction across the whole set: it depends on inheritance mode. In the 46 AD genes, the two cutoff sets were close on overall accuracy (90.6% derived against 89.7% panel-specified) yet split by class much as before. Derived pathogenic accuracy reached 100.0% against 98.0% for the panel-specified cutoffs, while derived benign accuracy fell to 34.8% against 57.1%. In the 27 AR genes, however, overall accuracy under the derived cutoffs was higher, reaching 92.4% against 86.9% for the panel-specified cutoffs, with derived benign accuracy (75.1%) now ahead of the panel-specified figure (58.0%) as well as derived pathogenic accuracy (98.7% against 98.3%). We had not expected the AD and AR comparisons to diverge this way.

The expert panel cutoffs are derived from disease-specific epidemiology through the Whiffin–Ware framework, which propagates prevalence, allelic heterogeneity, and penetrance into a maximum credible allele frequency. That approach is principled, but its inputs are hard to estimate reliably, and clinical prevalence estimates in particular tend to be drawn from ascertained cohorts that overrepresent severe presentations. When such estimates enter the calculation, the resulting benign thresholds can sit at lower frequencies than the empirical variant distribution supports, which is consistent with the drop in pathogenic-to-benign errors under the derived cutoffs. It does not, on its own, explain why the derived cutoffs called a larger share of benign variants incorrectly than the panel-specified cutoffs did on the AD genes specifically. This pattern does not hold in the AR genes, where the derived cutoffs called benign variants more accurately than the panels did. We cannot resolve either half of that split from these data alone.

The expert panels remain the appropriate reference where they exist; their per-gene curation incorporates evidence far beyond frequency. What the AD/AR split does suggest is that for the AF-based criteria specifically, thresholds fit directly to the empirical variant distribution can differ from those derived through the Whiffin–Ware pipeline in the direction the underlying input-estimation uncertainty would predict. On genes where the two agree, the derived tables provide independent support for the panel cutoffs. On genes where they disagree, the disagreement points to epidemiological inputs that may bear re-examination. For the approximately 99% of clinical genes that no VCEP currently covers, the derived cutoffs provide an alternative to fixed defaults grounded in the same calibration framework the panels apply on the covered genes. Where a panel specification exists, that specification is the appropriate primary reference and the derived cutoffs are intended as a complement, not a substitute.

### 4.3. Limitations

When filtering variants to build the dataset, we faced two options for variants absent from gnomAD: impute an allele frequency for them, or omit them entirely. A variant missing from gnomAD implies a true frequency somewhere between the minimum observable gnomAD AF and a singleton in the global population, a range spanning several orders of magnitude, which makes any imputation choice difficult to defend. To keep the analysis anchored in observed data we omitted these variants rather than assigning them pseudocount frequencies. The dropped variants are 3:1 pathogenic to benign, so the omission falls disproportionately on the pathogenic class. This bias is expected to attenuate over time: every human genome carries roughly 70 *de novo* variants, most of them benign, and as such variants accumulate in ClinVar across successive releases the proportion of benign variants absent from gnomAD will grow, narrowing the imbalance among dropped variants. The ClinVar labels this analysis treats as ground truth are not fully independent of the AF values against which the cutoffs are calibrated. Submitters classifying a variant under the ACMG/AMP framework may already apply PM2, BS1, or BA1, so the labels contain some contribution from the same evidence line the derived cutoffs seek to quantify. Frequency is 5 of the 28 ACMG/AMP criteria^1^, and a variant reaching a definitive classification rarely does so on frequency alone, so the effect is bounded but not zero. Its magnitude cannot be quantified from these data.

Several methodological choices constrain the scope and interpretation of the derived cutoffs. We restricted the calibration to missense variants in genes with autosomal dominant or autosomal recessive inheritance; loss-of-function variants, indels, X-linked genes, and mitochondrial genes are outside its remit and will require separate treatment as future work. We fixed the Bayesian prior probability of pathogenicity at 0.0441, the Pejaver 2022 point estimate^3^, and did not perform sensitivity analysis against alternative priors; cutoffs at strengths near the Bayesian evidence-strength boundaries could shift under a different choice. The stratification axis, missense O/E upper confidence bound, conflates gene-level constraint with gene length, since the upper confidence interval is mechanically wider for shorter genes; a short but genuinely constrained gene may thus appear less constrained than it is. The gnomAD exclusion falls hardest on the AD stratum, where 88.1% of pathogenic variants were removed, leaving an AD calibration set that is 6.8% pathogenic; PM2_Supporting sits nearest the resulting frequency floor and is the least well supported of the derived cutoffs. Finally, the VCEP comparison rests on 73 genes with parseable, gene-specific specifications available in the CSpec Registry and a purely autosomal dominant or recessive inheritance pattern, drawn largely from cardiac, cancer predisposition, and metabolic panels. This is a convenience sample rather than a representative cross-section of clinical genes, and generalization to the broader gene space cannot be assumed from these comparisons alone.

## Data Availability

All datasets used in this manuscript are publicly available for download.
All scripts used in this manuscript, and the resulting MOI $\times$ missense O/E cutoffs, are available in the Bitscopic BIAS-2015 repository, release v3.1.0 (2026-09-01, commit 9d8005b): \url{https://github.com/bitscopic/BIAS-2015/releases/tag/v3.1.0}.

https://github.com/bitscopic/BIAS-2015/releases/tag/v3.1.0

## Author Contributions

V.D., conceptualization, methodology, software, formal analysis, writing, review and editing C.E., conceptualization, supervision, review and editing

## Data and Code Availability

All datasets used in this manuscript are publicly available for download. All scripts used in this manuscript, and the resulting MOI *×* missense O/E cutoffs, are available in the Bitscopic BIAS-2015 repository, release v3.1.0 (2026-09-01, commit 9d8005b): https://github.com/bitscopic/BIAS-2015/releases/tag/v3.1.0.

